# The Peruvian Genome Project: expanding the global pool of genome diversity from South America

**DOI:** 10.1101/2024.05.05.24306840

**Authors:** Heinner Guio, Omar Caceres, Cesar Sanchez, Carlos Padilla, Omar Trujillo, Victor Borda, Luis Jaramillo-Valverde, Julio A. Poterico, Carolina Silva-Carvalho, Mary Horton, Cristina M Lanata, Alessandra Carnevale, Sandra Romero-Hidalgo, Samuel Canizales-Quinteros, Víctor Acuña-Alonzo, Marco Machacuay-Romero, Pedro Novoa, Roberto Frisancho, Ruth Shady, Pedro Flores-Villanueva, Timothy D. O’Connor, Manuel Corpas, Eduardo Tarazona-Santos

## Abstract

The process of inhabiting the Americas by ancestral native American populations involved many individuals settling in the Peruvian Andes and Amazonian regions. Due to Latin American countries representing less than 1% of the human genome data available in public reference databases, the evolution and migration processes involved in adapting to this unique geography have not yet been fully explained. The Peruvian Genome Project is an initiative, started in 2011, to address the underrepresentation of genomic data from native South American populations. This project has collected 1,149 samples from 17 traditional native and 13 mestizo (mixed of native, African, and European ancestry) communities. Currently, 150 whole genomes and 873 array-genotyped individuals have been sequenced from across the geography of Western South America, including coastal, Andes, and Amazonian regions. We discovered 1.6 million novel genetic variants with varying frequencies, indicative of local environmental adaptations and population drift. These novel variants allow us to infer local evolutionary traits and population-specific allele frequencies for people living at different altitudes, as well as varying adaptations to pathogens and living conditions. The Peruvian Genome Project is the result of over a decade of work in sample selection, logistics, and approved regulatory community engagement, designed to enhance the human genome pool of diversity of native Americans. The data collected here enable the targeted characterization of endemic diseases, trait adaptations, and new variants of clinical significance in South America. The Peruvian Genome Project represents a step forward in international and multidisciplinary efforts to make precision medicine more inclusive and accessible for underrepresented communities in Latin America, offering significant potential for drug development and diagnostics in a neglected continent.

## INTRODUCTION

Estimates of native American representation within genome reference datasets currently fluctuate between 1-5%, depending on the data source^1,2^. Peru, which encompasses an area of 1,285,215 km^2^, boasts a highly diverse population of 33 million inhabitants as of 2023, featuring both mestizos and native peoples, the two ethnic categories broadly used in Latin American societies and census^3^. Historical records indicate a significant demographic shift among its constituent native population: in 1620 (almost 90 years after the arrival of the first European conquerors), they amounted to 75% of Peru’s population, a figure which decreased to 56% by 1796 and further dwindled to 31% by 2003, reflecting in part a complex social process of urbanization of the Peruvian society. Today, native populations are fragmented into 47 distinct groups^4,5^, mostly rural, many of which have been inaccessible for genome and phenotypic characterization. The existing downward proportion trend underscores a concerning reality: the proportion of population considered as native is declining over time. This decline, due to colonization and migration, is not only steadily reducing opportunities for exploring the original genetic landscape of the Americas but also decreasing chances for charting endemic evolutionary adaptations specific to a unique set to climates and environments. Therefore, the observed decline in the population classified as native Peruvians translates into a continual reduction in the prospects for investigating novel genetic variants harbored by these communities. This is not only concerning for the local populations, as these variants explain specific characteristics relevant to genome medicine. It is also critical for the scientific community to act now and ethically engage with these understudied communities before the current window of opportunity disappears.

Some high Andean populations reside above 2500 meters above sea level (m.a.s.l.), which constitutes approximately 30% of the population of Peru^6^. The Andes and the Amazon jungle have influenced the environmental exposure of local populations. The population structure within the Western South America region has also exhibited remarkable homogeneity between Andean populations, suggesting a tendency to migrate within the Andean region^7^. This pattern of migration has likely conditioned populations to remain in their environment, fostering diverse adaptive changes, notably in response to hypoxia^8^. However, underlying genetic mechanisms for local adaptations, as well as their interactions with environmental factors, lifestyle, and potential epigenomic modifications, remain to be fully elucidated. These factors collectively contribute to physiological, endocrine, cardiovascular, respiratory, and other systemic changes observed in native populations^9,10^.

When carrying out the analysis of coastal, Andean, and Amazonian populations, it is useful to subdivide them according to their proportion of admixture. The examination of both mestizo and native population classification offers a valuable framework for advancing genomic medicine in Peru and the broader Americas^11,12^ and has the potential to delineate health disparities and disease prevalence in these communities. While scientific discoveries are starting to emerge from the Peruvian Genome Project (PGP), it is essential to acknowledge its limitations, which mostly relate to its modest size. Nevertheless, these limitations are significantly offset by the stringent sample selection criteria with which sampling diversity was performed and the extraordinary diversity offered by some populations, which as of to date have not been studied before. These project findings, as illustrated in **Figure 1**, pave the way for further advancements toward precision medicine and data equity within a dramatically underrepresented set of human populations in the Latin American region

**Figure 1:**
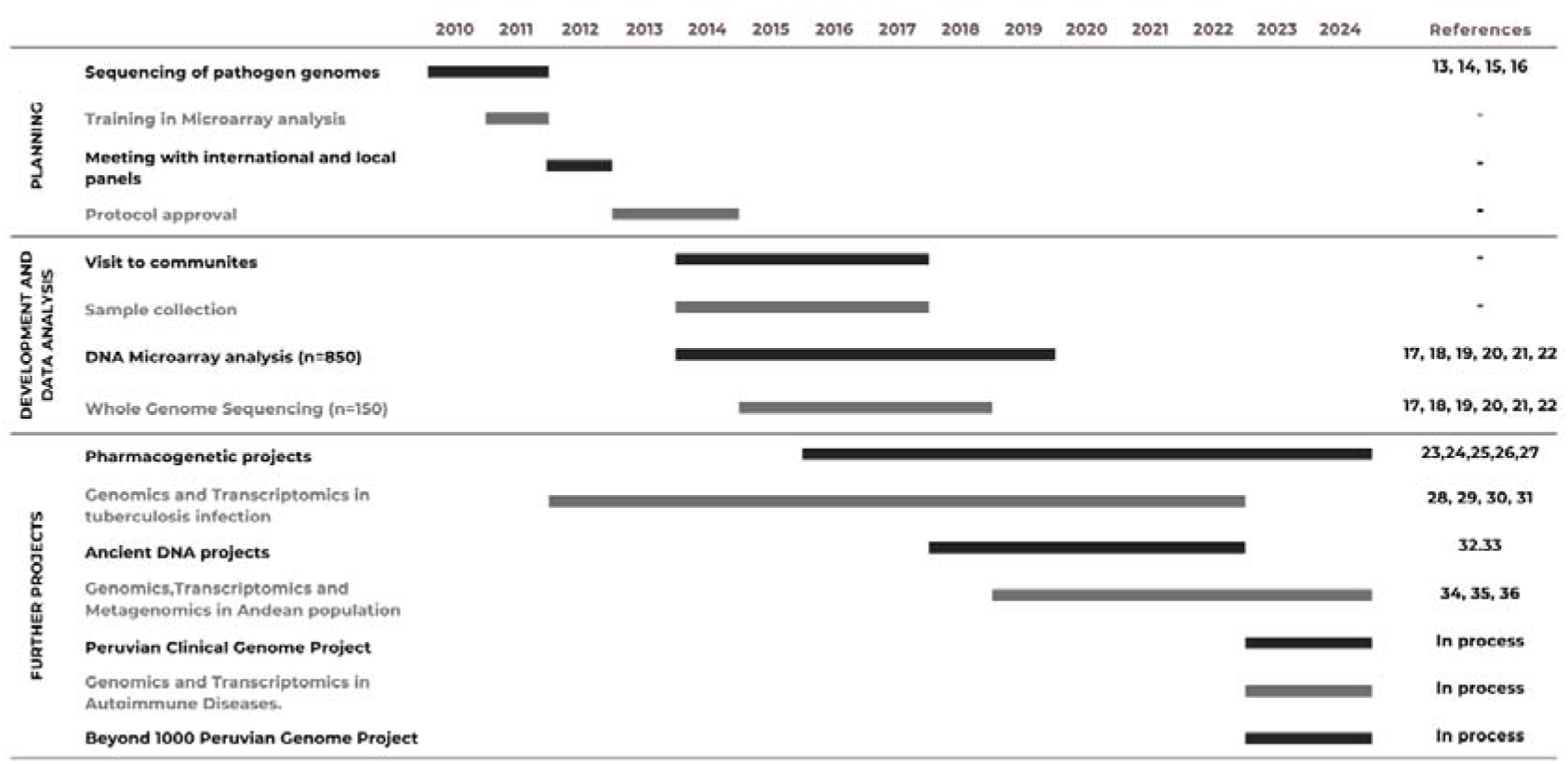
Timeline for the development of the Peruvian genome project and future perspectives.

The initial stages of the PGP involved the sequencing of pathogen genomics in 2010. Since then, the project evolved into a tailored-made protocol for selecting populations and individuals for sequencing and genotyping from across the diverse geography of Western South America. This protocol involved international experts and local authority panels, starting in 2012. It took us over a year to have a suitable ethical and sample logistics procedure approved by regulatory authorities. In 2013, we then started visiting communities with which we engaged via a variety of channels, including local media, presentations, and interviews. These interviews included chiefs (“Apus”) from local communities as our initial point of contact and exchange of information. Once appropriate consent from Apus and authorized individuals was obtained, sample collection, sequencing, and genotyping started in 2014 and went through to 2017. The sequencing was performed at the New York Genome Center (NYGC) and the array genotyping at the facility of the Instituto Nacional de Salud del Peru [Peruvian National Institute of Health]. It took us 3 years for this phase to be completed, including the implementation of anthropological and genetic analyses, which culminated with the first peer-reviewed publication of initial results in 2018^22^. Herein, we showed that the studied populations of the three geographic regions in Peru (Amazon, Andes, and coast) diverged from each other ∼12,000 ya. In 2018 and 2019 we then performed pharmacogenomics and transcriptomics investigations. This research allowed us to shed light on the susceptibility’s highlanders have in tuberculosis infections, together with their pharmacological responses^23, 28^. During the COVID pandemic (2020-2022), we shifted our focus to collaborating with international research groups to incorporate ancient Peruvian DNA samples into the pool. From then onwards, we focused our efforts on developing a pool of genome and phenome data that goes beyond the current ∼1000 genomes we have collected to date that are in the process of sequencing. As we continue exploring the wealth in diversity that the Andes, Amazon, and Coast accumulate in Western South America, we have worked with special emphasis on the endemic diseases and precision medicine approaches that apply to local native populations.

## POPULATION SELECTION CRITERIA AND SAMPLE COLLECTION STRATEGIES

The presence of the Andes, which cross from North to South the South American continent **(Figure 2)**, has dramatically shaped human evolution and adaptation in the region. Nearly 30% of the Peruvian population live above 2,500 m.a.s.l. on the Andes, whereas 10% live in the Amazon jungle, on the East side of the mountains, and all of them exhibit different epidemiological and isolating environmental conditions^37^. As indicated above, to carry out a balanced recruitment and identification of population diversity criteria by the mandate of the Instituto de Salud Nacional del Perú, we convened a local and an international research panel as follows. **i) The Local Panel** was composed of representatives of the native communities of Peru, the Ministerio de Cultura, Non-Governmental Organizations, archaeologists, sociologists, and anthropologists. According to the experience of this panel, and the main objective of the project, 3 criteria were identified to select populations from across the coast, Andes, and Amazon jungle: representativeness (number of residents in each population), degree of isolation, and vulnerability of the population to extinction. Within these criteria, 17 native populations and 13 mestizo populations were identified, spanning diverse locations and geographical distances (**Figure 2**). **Figure 3** shows the total breakdown of community participant individuals from a total of 1,149 individuals. Additionally, for this process of selection we also considered migratory routes and self-described identity information of populations to understand historical movements of peoples across centuries and processes of miscegenation within the communities. We applied a definition of “native individuals” consisting of parents and grandparents of the subject under study being born in the same native community. **ii) The International Panel** was composed of researchers who had already developed projects in similarly underrepresented countries, such as researchers from INMEGEN (National Institute of Genomic Medicine of Mexico), the University of Michigan, University of Maryland (USA), and Universidade Federal de Minas Gerais (Brazil). These researchers’ experience allowed us to consider appropriate clinical measurements and analyses not necessarily considered in previously peer-reviewed projects.

**Figure 2:**
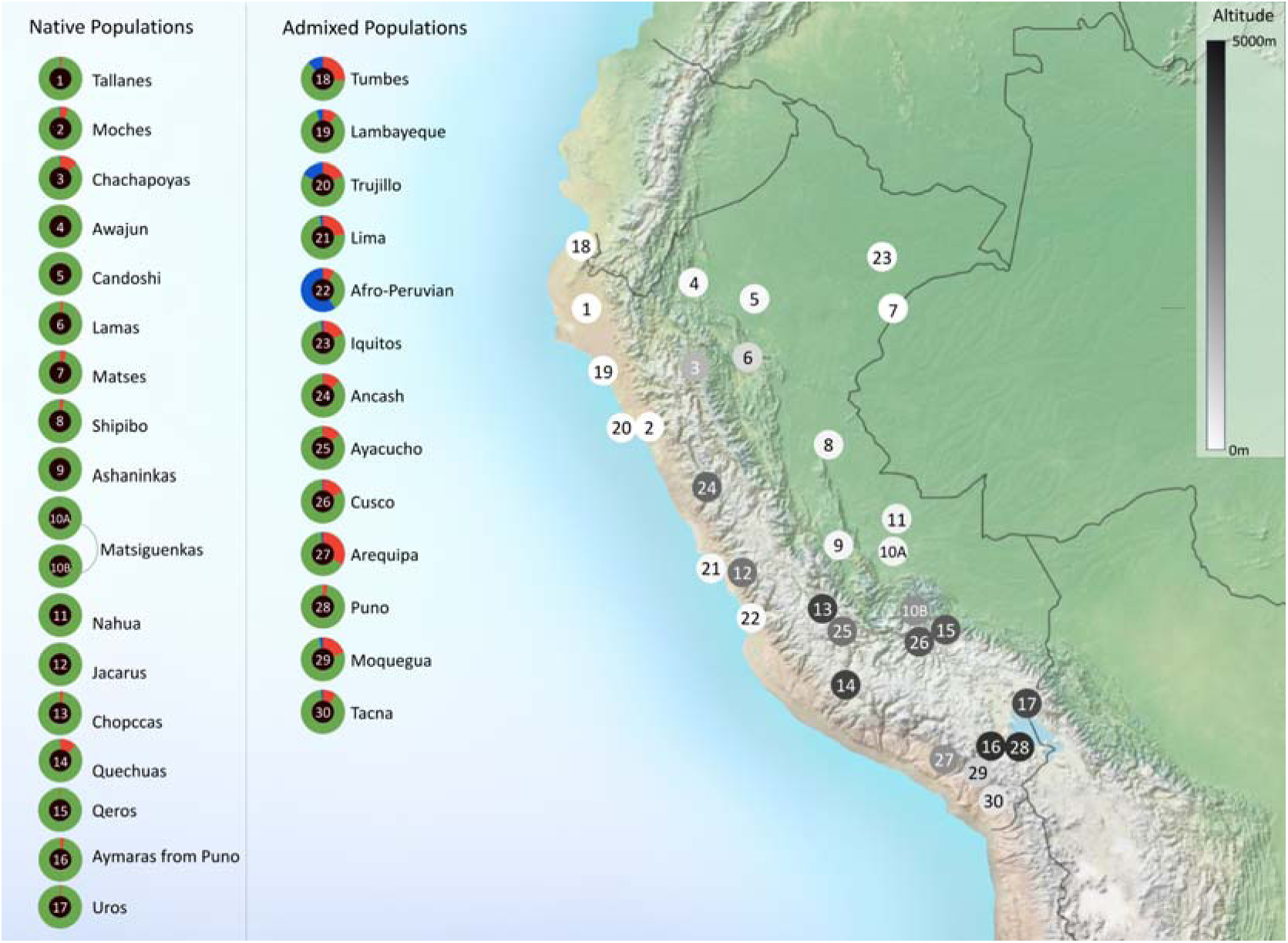
Genome-wide ancestry proportions and geographic distribution of Peruvian populations selected for the Peruvian Genome Project. Doughnut plots show the degree of admixture for each population, inferred by the mean of the result of K=3 from the ADMIXTURE software^38^. The ancestry components of African, European, and Native American origins are represented within these doughnuts in blue, red, and green, respectively. An adjacent map shows the location of the different populations in numbered circles shaded with different intensities to indicate the average altitude at which the population usually resides.

**Figure 3:**
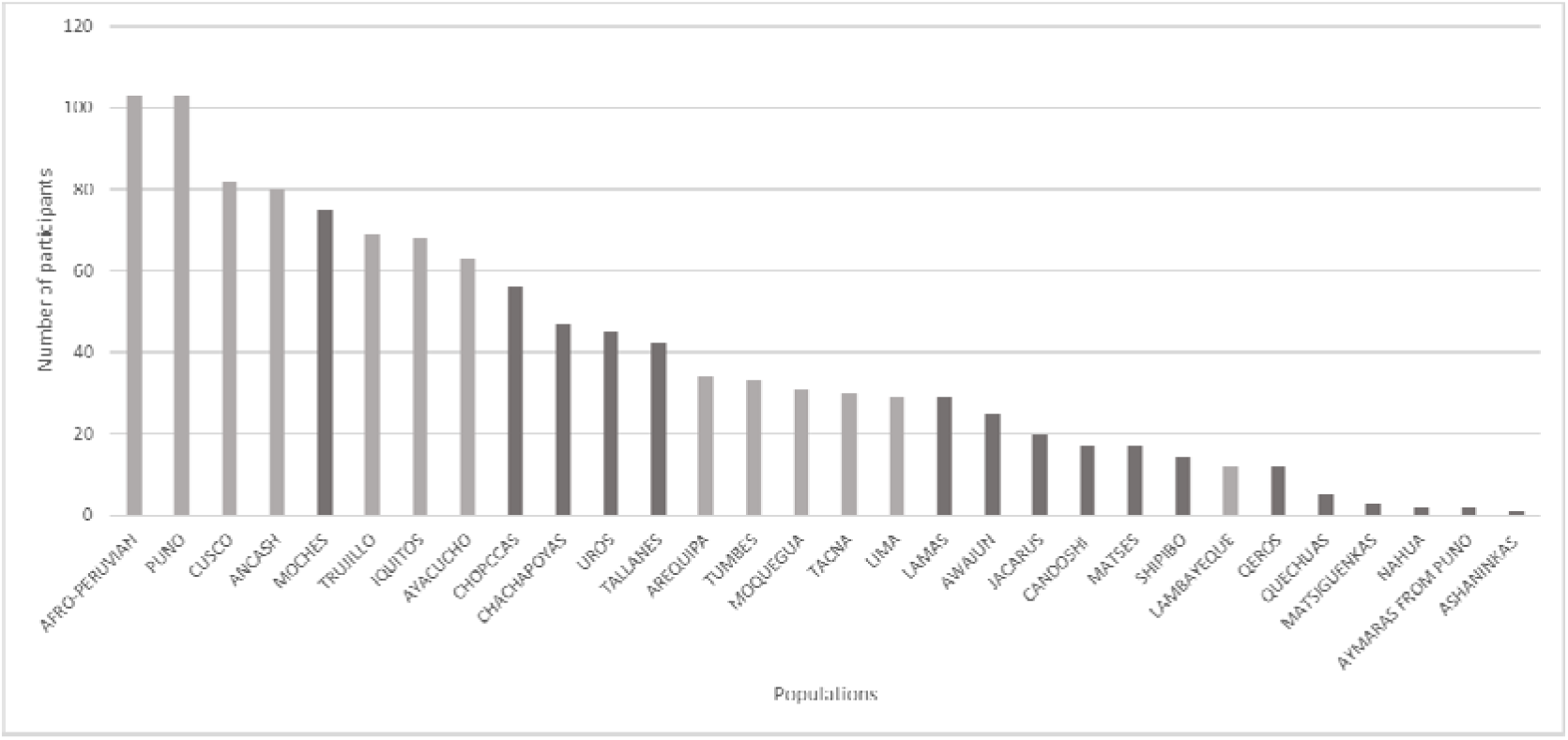
Total number of participants recruited for the Peruvian Genome Project (n=1,149) and the breakdown of communities from which they come from black: native populations, grey: admixed population.

## ETHICAL APPROVAL PROCESS AND CASCADE OF CONSENTS

The ethical process for collection, stewardship, and dissemination of data and results for gathered samples was meticulously implemented and carried out at the community and individual levels. We made sure that communities and individuals were engaged according to international and local protocols, including the Declaration of Helsinki for medical research involving human subjects. For community participation, a consultation process was performed involving authorities at the national level (Ministerio de Salud, Peruvian Government), regional authorities, and several Peruvian universities. The information gathering strategy for sample collection began a month in advance for each community, including communication materials. These materials consisted of: i) the distribution of a brochure that explains the project written in simple language (Spanish or local native language); ii) the display of a poster that reproduces the informed consent format; iii) public informative sessions aimed at the participating community; and iv) communication through local television, radio, and written press whenever possible or available. Native communities were visited several months before sampling to request authorization from the community leaders (Apus). The final decision for participation was made by the individuals themselves who had to understand and consent to the ethical processes outlined here. Subjects that matched the inclusion criteria were contacted to participate and then informed consent and authorization were obtained to preserve their samples. All participants were offered the possibility of withdrawing from this study at any time with no need for explanation required. All participants gave their informed consent in the presence of a translator to their mother-tongue traditional language and two local witnesses. All procedures were reported and presented for evaluation and approval by the Research and Ethics Committee from the Instituto Nacional de Salud (authorization no. OI-003-11 and no. OI-087-13).

## DATA SAMPLING AND GENOTYPING

Participants were selected to represent diverse self-described Native and Mestizo Peruvian populations. We applied three criteria to optimize participant selection to best represent the Indigenous American populations. These included: (i) the place of birth of the participant, his or her parents and grandparents (they all had to belong to the same community), (ii) their last names (selecting only those corresponding to the region if they existed or were known), and (iii) if several members of a family met our standards for inclusion, the oldest member was then selected for our study.

The first phase of this study included 150 Native and Mestizo Peruvian whole genomes, sequenced to an average of 35X coverage on an Illumina HiSeq X 10 platform by the New York Genome Center (NYGC). An additional 130 Native American and mestizo Peruvian individuals were genotyped using a 2.5M Illumina chip, featuring over two-millions of markers for dense genome-wide coverage and extensive disease-associated content at the Biotechnology and Molecular Biology Lab of the “Instituto Nacional de Salud del Perú”.

## PHENOTYPE DATA AND LABORATORY MEASUREMENTS

To date, we have collected a total of 1,149 samples (each corresponding to a different individual) from 17 traditional Native and 13 Mestizo (admixed of Native Peruvian, African, Asian, and European ancestry) communities.

Demographic information, including lifestyle (smoking status and diet patterns), and anthropometrics data such as body mass index (BMI), weight, and height were measured. These measurements were also included in fasting conditions to encompass blood lipids, blood cell traits (mean hemoglobin levels, red cell count, white cell count, and platelets), glucose levels, and renal function markers (Table 1).

**Table 1.** Characteristics of participants enrolled in the Peruvian Genome Project (PGP).

| Characteristic | All (n=1149) | Native (n= 506 ) | Mestizo (n=643 ) |
| --- | --- | --- | --- |
| Number of individuals interviewed, N | 1149 | 506 | 643 |
| Gender N(%) |  |  |  |
| Female | 382 (33.3) | 185 (36.6) | 197 (30.6) |
| Male | 767 (66.7) | 321 (63.4) | 446 (69.4) |
| Location |  |  |  |
| North | 229 (22.8) | 114 (24.5) | 115 (24.5) |
| South | 194 (19.3) | 16 (3.4) | 178 (38.0) |
| Center | 354 (35.3) | 178 (38.2) | 176 (37.5) |
| East | 226 (22.6) | 158 (33.9) | 0 (0.0) |
| Age (years), median (IQR) | 41 (23-56) | 53 (38-68) | 32 (21-41) |
| Age group (years), N (%) |  |  |  |
| 18-19 | 98 (8.7) | 7 (1.4) | 91 (14.2) |
| 20-29 | 363 (32.3) | 60 (12.3) | 303 (47.4) |
| 30-39 | 142 (12.6) | 68 (14.0) | 74 (11.6) |
| 40-49 | 163 (14.5) | 90 (18.5) | 73 (11.4) |
| 50-59 | 142 (12.6) | 87 (17.9) | 55 (8.6) |
| 60+ | 217 (19.3) | 174 (35.8) | 43 (6.7) |
| Body mass index (kg/m<sup>2</sup>), mean ± SD | 25.4 ± 5.58 | 26.0 ± 5.19 | 24.9 ± 5.83 |
| BMI classification, N (%) |  |  |  |
| Underweight | 25 (2) | 12 (2) | 13 (2) |
| Normal | 516 (45) | 198 (39) | 318 (50) |
| Overweight | 423 (37) | 198 (39) | 255 (35) |
| Obese | 177 (16) | 96 (20) | 81 (13) |
| Smoking status, N (%) |  |  |  |
| Current smoker | 139 (12.1) | 64 (12.7) | 75 (11.7) |
| Never smoked | 1006 (87.9) | 441 (87.3) | 565 (88.3) |
| Alcohol consumption, N (%) |  |  |  |
| Never or no alcohol use | 647 (56.8) | 306 (60.8) | 341 (53.6) |
| Frequent current drinker | 493 (43.2) | 197 (39.2) | 295 (46.4) |
| Metabolic markers (mean ± SD) | | | |
| Total cholesterol (mg/dL), mean ± SD | 163.1 ± 48.18 | 169.0 ± 50.83 | 159.4 ± 46.08 |
| High-density lipoprotein (mg/dL), mean ± SD | 53.0 ± 19.06 | 55.5 ± 18.55 | 49.0 ± 19.20 |
| Low-density lipoprotein (mg/dL), mean ± SD | 85.7 ± 50.14 | 93.8 ± 53.17 | 80.7 ± 47.51 |
| Triglycerides (mmol/L), mean ± SD | 127.8 ± 83.59 | 138.7 ± 88.15 | 120.9 ± 79.89 |
| Bilirubin, median (IQR) | 0.8 (0.56-1.30) | 1.0 (0.72-1.6) | 0.8 (0.48-1.11) |
| Blood Pressure |  |  |  |
| Systolic pressure (mmHg), mean ± SD | 111.9 ± 18.5 | 115.5 ± 22.8 | 109.6 ± 14.7 |
| Diastolic pressure (mmHg), mean ± SD | 69.7 ± 48.18 | 69.9 ± 12.3 | 69.5 ± 10.0 |
| Anthropometric Measurements |  |  |  |
| Weight (kg), mean ± SD | 63.7 ± 14.61 | 63.2 ± 14.26 | 64.1 ± 14.88 |
| Height (cm), mean ± SD | 157.3 ± 9.09 | 155.0 ± 9.30 | 159.1 ± 8.49 |
| Anemia |  |  |  |
| White blood cell count | 8.1 ± 4.51 | 8.1 ± 5.46 | 8.1 ± 3.81 |
| Red blood cell count | 45.3 ± 13.77 | 43.6 ± 6.95 | 46.4 ± 16.59 |
| Hemoglobin | 14.9 ± 2.09 | 14.4 ± 2.33 | 15.18 ± 1.87 |
| Platelet count (PLT) | 316.9 ± 76.94 | 296.4 ± 81.09 | 329.6 ± 71.40 |
| Lymphocytes | 29.3 ± 11.80 | 28.2 ± 14.14 | 29.9 ± 10.02 |
| Monocytes | 4.1 ± 2.94 | 4.4 ± 3.20 | 3.9 ± 2.75 |
| Basophils | 0.2 ± 0.61 | 0.2 ± 0.59 | 0.3 ± 0.62 |
| Eosinophils | 2.7 ± 3.72 | 3.6 ± 5.05 | 2.2 ± 2.42 |

We were able to infer significant differences between Native and Mestizo samples due to environmental factors such as geographical altitude, and diet. BMI and ancestry, which have been associated with specific environmental conditions in previously related studies^39^, showed significant differences in BMI between Native and Mestizo populations. However, more precise methodologies need to be considered in future studies in order to discriminate whether these differences are related to excess fat or muscle mass percentage.

## INITIAL FINDINGS AND KEY CONTRIBUTIONS FOR GENOMIC MEDICINE IN PERU AND NATIVE AMERICANS

The PGP offers significant insights into the genetic diversity and evolutionary adaptations of Native and Mestizo populations in Peru. To date, PGP has sequenced 150 genomes and genotyped 850 individual samples. This work is being applied to a range of clinical interventions that are actionable for the advancement of precision medicine, public health strategies, and understanding of human genetic evolution^40^. In what follows, we present some examples that illustrate how our work has shed light on our understanding of native Peruvian genome variants endemic from these populations.

### Enhanced Understanding of Genetic Diversity and Disease Susceptibility

It has been previously shown that the Peruvian Mestizo populations have at least a 60% genetic Indigenous American ancestry, with some native communities adding up to 90% of their genetic native component as shown in Figure 2. We have discovered 1.6 million novel genetic variants according to the Variant Effect Predictor (https://grch37.ensembl.org/Homo_sapiens/Tools/VEP), which are not present in existing data banks resources such as dbSNP^13^. The inclusion of variants from under-represented populations such as Native Peruvians in global genomic databases is expected to aid in refining and broaden the accuracy of genetic risk assessments and pharmacogenomics interventions for diseases most prevalent for indigenous American populations, whose pharmacogenomic representation is only 0.1% of existing data^41^. These efforts will then lead to more personalized and effective healthcare interventions, at least to some degree, for all Indigenous American groups.

One of the initiatives preceding the Peruvian Genome Project was the 1000 Genomes Project^42^, which included 85 genomes of Peruvians from Lima. These individuals, although helpful, were all sampled from Mestizo individuals from Lima, so have more European ancestry were not representative of the diversity richness of Native communities in Peru. The advantage of the PGP lies in the sophisticated inclusion criteria carried out to consider different biogeographical regions from across the coast, the Andes, and the Amazon. This allowed us to evaluate the effects of migration over thousands of years as well as helping differentiate populations based on evolutionary bottlenecks, revealing a distinct genetic fingerprint on the Americans’ ancestors^43^. Moreover, we have found that the degree of ancestry (mestizos vs natives) and the geographic altitude habitat (high vs low landers) are linked^23,34^.

Unique genomic changes in the composition of Peruvian populations, such as the ones we are beginning to uncover, have aided in elucidating the impact of ancient migrations, helping determine population structure based on geographic barriers in Peru^34,44,45^. Our data provides evidence of migrations in the central and northern Andean and Amazon regions, which are restricted in the Southern Andes and are likely due to the effect of the high elevation of the Andean mountains. Evidence of gene flow from migrations and differential patterns of genetic variation have also been found, including those associated with immune system genes^23^.

### Pharmacogenomics and Drug Safety

Genetic ancestry plays a central role in population pharmacogenomics^46^. In one of our studies, we researched the presence of adverse reactions during anti-tuberculosis treatment in the Peruvian population. Our results suggest that 30% of the Peruvian populations are associated with the slow metabolism of isoniazid^24^. We also identified haplotypes with divergent associations with drug-induced liver injury (DILI), based on the mestizo or native Peruvian population. For instance, we found evidence of *NAT25B* and *NAT2*7B being associated with DILI risk in mestizos, while no such association has been observed in natives. Additionally, haplotypes *NAT25G* and *NAT2*13A have only been negatively associated with DILI in the studied Native Peruvians^23^. Current research also suggests a greater prevalence of probable hepatotoxicity in the Amazonian population. In a study still in progress, we have compared the pharmacogenetic response to antituberculosis drugs in a population with ancestry greater than 95% native from the coast, Andes, and the Amazon of Peru. Our initial results point to greater hepatotoxicity in antiretroviral cotreatment on the coast. To exemplify the importance of more human genomics studies on underrepresented populations, we consider important to identify SNVs clinically relevant in pharmacogenetics (levels 1 and 2 of the PharmGKB database) that could show differences between the Andean and Amazonian populations. Thus, by considering the genetic diversity within and between populations, healthcare providers are beginning to better predict adverse drug reactions, adjust dosages, and select the most appropriate medication, thereby enhancing patient safety and treatment efficacy^25,26^.

### Adaptation to High Altitude and Its Health Implications

PGP’s findings on the selection of genes related to immune response in different Peruvian populations offer promising data for understanding susceptibility and resistance to infectious diseases. We revealed that this environmental and genetic differentiation between the Andean and Amazonian populations has allowed natural selection and other evolutionary forces to act over millennia, shaping differences in the frequencies of genetic variants, including genes related to the immune response (*CD45* and *DUOX2*), with thyroid (*DUOX2*), cardiovascular (*HAND2-AS1*) and hematological (*TMPRSS6*) functions 4, as well as genes related to drug response^34^. Considering that bottleneck effects and genetic fixations have shaped the biological structure marked by geographical differences in Peru, our group embarked on finding how this effect may be manifested in gene expression.

In previous studies, the immune response has been mainly associated with ancestry. However, our findings show that altitude and the microbiome are also as important as ancestry in the immune response in populations with a high Indigenous American ancestry. To evaluate the immune response in Native Highlanders, a laboratory was developed at the University of Huanuco, located at 3000 meters above sea level, to carry out a transcriptomic project resulting from the stimulation of PBMCs with proteins from bacteria, viruses, and fungi. For this aim, we intended to address the research question of how the expression of immune response-associated genes varies in high-altitude populations compared to those not inhabiting such geographic conditions. Our unpublished data suggest that there is indeed a difference in the immune response of these native inhabitants, with upregulated expression in genes such as *HLA-DPB1, FN1, CD36, MMP2, HLA-DRB1, FCGR1A, CCL17, and HLA-DRB5*, and down-regulation in *TGAX, CCL22, CSF1, CXCL8, IL12A, MMP9, CSF2, PTGS2*, and *FGF2*. However, at the genomic level, we have not found variation in gene expression in those genes associated with the evolutionary pattern of native populations in the central Peruvian highland region^35^. Our findings agree with recent observations by Sharma *et al.*, who suggested that the effects of genetic selection do not align with genes exhibiting higher or lower genetic expression, nor within the realm of proteins^47^.

### Prevalence of established autoimmune risk variants in the PGP

Autoimmune diseases, in particular Systemic Lupus Erythematosus (SLE), tend to be present at a younger age, and with worse clinical outcomes, in people of non-European descent. Immune genetic variants show differential evolution based on geographic pathogen pressure as nearly 13% of non-HLA GWAS loci for SLE exhibit signs of natural selection^48^.

While acknowledging the inherent challenges in comparing GRSs across diverse populations, in this case constructed from Genome-Wide Association Study (GWAS) data predominantly sourced from European and Asian cohorts, it’s noteworthy that healthy Native Peruvians exhibit elevated unweighted polygenic risk for SLE in contrast to European, African, and South Asian counterparts, aligning more closely with East Asians. Confirmation of these findings necessitates comprehensive population-based studies within the Americas. These findings emphasize the imperative for continued genetic investigations into autoimmunity in Peru, moving beyond Eurocentric genetic frameworks, and offering potential insights into the heterogeneity of SLE.

## NEXT STEPS

### Ancient DNA studies in samples from 8000 years ago

Genetic variants have been generated throughout human evolution and migration since the first inhabitants. Environmental factors, epidemiological pressures, and human interactions have likely played roles in shaping the emergence and persistence of genetic variants. However, the origins of these variants in the context of Peru’s ancient populations remain unclear. It is uncertain whether these variants were introduced by the first immigrants or emerged locally. Conducting ancient DNA studies in Peru holds the potential to illuminate this question.

For this purpose, we have generated a mobile laboratory installed in the excavation centers so after the identification of coprolites, samples are transported in 5 minutes to the lab to start extracting ancient DNA and avoid contemporary contamination^32,33^.

### Peruvian Clinical Genome

Previously, we found SNPs associated with the prognosis, severity and treatment response in tuberculosis infection^28^. As the pathogenic potential of newly discovered genetic variants within the PGP remains uncertain, a collaborative effort with the University of Westminster is underway to establish a comprehensive database of the Peruvian clinical genome. This database aims to enable and streamline specialized medical studies, elucidating the clinical significance of these variants in relation to Peru’s most prevalent diseases.

### Genomics and transcriptomic in cardiovascular-related genes in highlanders

Considering the new genetic variants associated with cardiovascular health found in the PGP, associated with the concept that not everything your genes have is expressed, we decided to combine genomic and transcriptomic analysis of cardiovascular genes in response to high altitude. This effort is in collaboration with Queen Mary University of London.

### Beyond 1000 Peruvian Genome Project (B1000PGP)

The rigorous data collection methodology adopted by PGP was developed in concordance with the *a priori* inquiries posed by an international panel of experts from related projects, specifically regarding opportunities and variables relevant to include in the PGP framework. We are currently deliberating the inception of B1000PGP, a project with heightened ambitions aimed at collecting a broader array of variables beyond genomic data, incorporating additional samples progressively over time. We are conducting assessments on both healthy individuals and those diagnosed with various pathologies. Our objectives encompass more than just elucidating the spectrum, prevalence, and implications of germline or tumor genomic variants. Moreover, acknowledging the pivotal roles of the microbiome and DNA methylation as essential markers in human health, we are deliberating additional initiatives to produce datasets of this type.

## CONCLUSIONS

Peru’s limited engagement in genomic research stems from a confluence of factors: foremost, the nation’s lack of prioritization of genomic medicine policies leads to a dearth of strategic direction and investment. Additionally, the scarcity of adequately trained genomic scientists impedes the development of a proficient workforce capable of advancing research in this field. Meanwhile, centralized research infrastructure in the capital city restricts access and opportunities for researchers in the countryside. To these difficulties, cumbersome administrative processes further hamper Peru’s ability to undertake large-scale genomic studies. Nevertheless, the PGP has managed to implement a multidisciplinary approach throughout years of work and international collaborations, establishing rigorous criteria for the selection and definition of mestizo and native populations. Ethical considerations and stringent methodologies, alongside strict inclusion criteria, have facilitated the identification of participants with substantial Indigenous American ancestry, yielding 1.6 million novel genetic variants pertinent to understanding migration, adaptation, and immune response in native Americans and highlanders. These findings bear great promise for potential clinical applications, yet discerning the biological adaptations of Peruvians remains paramount, as genomic variation devoid of clinical significance merely constitutes a discovery without necessarily leading to actionability. To propel genomic research forward in Peru, fostering international collaborations, particularly through training grants for doctoral and post-doctoral positions, is imperative. Equally crucial is the development of infrastructure conducive to initiating new projects and facilitating the interpretation of results. Such multidisciplinary collaborative efforts are indispensable for elucidating the genomic heritage of Peruvian populations residing in the Andean and Amazonian regions, thereby enriching our collective understanding of humankind.

## ACKNOWLEDGMENTS

To the researchers who were part of the national panels (Cesar Cabezas, Sonia Guillen, Oswaldo Salaverry, Santiago Pastor, Patricia Mayta, Leonidas Gomez, Evelyn Guevara) and Angel Medina, Yuri Alegre, Harrison Montejo for their invaluable support in the coordination and collection of samples. To Luis Guillermo Lumbreras for his advice on anthropological and archaeological topics. To facilitate studies in highlanders Jose Beraun, Milward Ubillus, Diana Palma. To all participants of this study.

## Data Availability

Data have been deposited in the European Genome-phenome Archive (EGA), https://www.ebi.ac.uk/ega/home (accession nos. EGAD00010001958, EGAD00010001990, EGAD00010001991, EGAD00010001992). Access to the data is available upon request of the authors through, please contact the corresponding authors for the access directions.

## INCLUSION AND DIVERSITY

We support inclusive, diverse, and equitable conduct of research.

## REFERENCES

1. Mills MC, Rahal C. The GWAS Diversity Monitor tracks diversity by disease in real time. Nat Genet. 2020 Mar;52(3):242–243. doi: 10.1038/s41588-020-0580-y.

2. Corpas M, Siddiqui MK, Soremekun O, Mathur R, Gill D, Fatumo S. Addressing Ancestry and Sex Bias in Pharmacogenomics. Annu Rev Pharmacol Toxicol. 2024 Jan 23;64:53–64. doi: 10.1146/annurev-pharmtox-030823-111731. Epub 2023 Jul 14. PMID: 37450899.

3. Salzano FM, Bortolini MC. The Evolution and Genetics of Latin American Populations. Cambridge: Cambridge University Press; 2001. (Cambridge Studies in Biological and Evolutionary Anthropology). 10.1017/CBO9780511666100

4. Le Gootenerg, P. (1995). Población y etnicidad en el Perú republicano (siglo XIX): algunas Revisiones. Documento de Trabajo, 71. Serie Historia, 14. http://repositorio.iep.org.pe/handle/IEP/994

5. Instituto Nacional de Estadística e Informática (2017). Perú: Perfil Sociodemográfico. Informe Nacional. Censos Nacionales 2017: XII de Población, VII de Vivienda y III de Comunidades Indígenas. Retrieved from https://www.inei.gob.pe/media/MenuRecursivo/publicaciones_digitales/Est/Lib1539/libro.pdf

6. Tremblay JC, Ainslie PN. Global and country-level estimates of human population at high altitude. Proc Natl Acad Sci. 2021 May 4;118(18):e2102463118.

7. Tarazona-Santos E, Carvalho-Silva DR, Pettener D, Luiselli D, De Stefano GF, Labarga CM, Rickards O, Tyler-Smith C, Pena SD, Santos FR. Genetic differentiation in South Amerindians is related to environmental and cultural diversity: evidence from the Y chromosome. Am J Hum Genet. 2001 Jun;68(6):1485–96. doi: 10.1086/320601. Epub 2001 May 15. PMID: 11353402; PMCID: PMC1226135.

8. Xing G, Qualls C, Huicho L, Rivera-Ch M, Stobdan T, Slessarev M, Prisman E, Ito S, Wu H, Norboo A, Dolma D, Kunzang M, Norboo T, Gamboa JL, Claydon VE, Fisher J, Zenebe G, Gebremedhin A, Hainsworth R, Verma A, Appenzeller O. Adaptation and mal-adaptation to ambient hypoxia; Andean, Ethiopian and Himalayan patterns. PLoS One. 2008 Jun 4;3(6):e2342. doi: 10.1371/journal.pone.0002342. Erratum in: PLoS ONE. 2008;3(6). doi: 10.1371/annotation/500fe2d3-85ad-47e2-a1a2-356a3125df68. Erratum in: PLoS ONE. 2008;3(6). doi: 10.1371/annotation/aba20dc1-b10d-464c-9671-c47b956d1718.

9. Bigham A, Bauchet M, Pinto D, Mao X, Akey JM, Mei R, et al. Identifying Signatures of Natural Selection in Tibetan and Andean Populations Using Dense Genome Scan Data. Begun DJ, editor. PLoS Genet. 2010 Sep 9;6(9):e1001116.

10. Beall CM. Tibetan and Andean patterns of adaptation to high-altitude hypoxia. Hum Biol. 2000 Feb;72(1):201–28. PMID: 10721618.

11. Tishkoff SA, Verrelli BC. Patterns of human genetic diversity: implications for human evolutionary history and disease. Annu Rev Genomics Hum Genet. 2003;4:293–340. doi: 10.1146/annurev.genom.4.070802.110226. PMID: 14527305.

12. Jorde LB, Watkins WS, Bamshad MJ. Population genomics: a bridge from evolutionary history to genetic medicine. Hum Mol Genet. 2001 Oct 1;10(20):2199–207. doi: 10.1093/hmg/10.20.2199. PMID: 11673402.

13. Tarazona D, Borda V, Galarza M, Agapito JC, Guio H 2014. Functional Analysis Using Whole-Genome Sequencing of a Drug-Sensitive Mycobacterium tuberculosis Strain from Peru. Genome Announc 2:10.1128/genomea.00087-14.

14. Galarza M, Tarazona D, Borda V, Agapito JC, Guio H 2014. Evidence of Clonal Expansion in the Genome of a Multidrug-Resistant Mycobacterium tuberculosis Clinical Isolate from Peru. Genome Announc 2:10.1128/genomea.00089-14.

15. Guio H, Tarazona D, Galarza M, Borda V, Curitomay R. 2014. Genome Analysis of 17 Extensively Drug-Resistant Strains Reveals New Potential Mutations for Resistance. Genome Announc 2:10.1128/genomea.00759-14.

16. Tarazona D, Galarza M, Levano KS, Guio H. [Comparative genomic analysis of peruvian strains of Mycobacterium tuberculosis]. Revista Peruana de Medicina Experimental y Salud Publica. 2016 Jun;33(2):256–263. PMID: 27656924.

17. Asgari, S., Luo, Y., Akbari, A. et al. A positively selected FBN1 missense variant reduces height in Peruvian individuals. Nature 582, 234–239 (2020). 10.1038/s41586-020-2302-0

18. Maíra R Rodrigues, Fernanda Rodrigues-Soares, Hanaisa P Sant Anna, Meddly L Santolalla, Marília O Scliar, Giordano Soares-Souza, Roxana Zamudio, Camila Zolini, Maria Catira Bortolini, Michael Dean, Robert H Gilman, Heinner Guio, Jorge Rocha, Alexandre C Pereira, Mauricio L Barreto, Bernardo L Horta, Maria F Lima-Costa, Sam M Mbulaiteye, Stephen J Chanock, Sarah A Tishkoff, Meredith Yeager, Eduardo Tarazona-Santos, Origins, Admixture Dynamics, and Homogenization of the African Gene Pool in the Americas, Molecular Biology and Evolution, Volume 37, Issue 6, June 2020, Pages 1647–1656, 10.1093/molbev/msaa033

19. Guio H, Poterico JA, Levano KS, Cornejo-Olivas M, Mazzetti P, Manassero-Morales G, Ugarte-Gil MF, Acevedo-Vásquez E, Dueñas-Roque M, Piscoya A, Fujita R, Sanchez C, Casavilca-Zambrano S, Jaramillo-Valverde L, Sullcahuaman-Allende Y, Iglesias-Pedraz JM, Abarca-Barriga H. Genetics and genomics in Peru: Clinical and research perspective. Mol Genet Genomic Med. 2018 Nov;6(6):873–886. doi: 10.1002/mgg3.533. PMID: 30584990; PMCID: PMC6305655.

20. Naslavsky, M.S., Scliar, M.O., Yamamoto, G.L. et al. Whole-genome sequencing of 1,171 elderly admixed individuals from Brazil. Nat Commun 13, 1004 (2022). 10.1038/s41467-022-28648-3

21. Victor Borda, Douglas P. Loesch, Bing Guo, Roland Laboulaye, Diego Veliz-Otani, Jennifer N. French-Kwawu, Thiago Peixoto Leal, Stephanie M. Gogarten, Sunday Ikpe, Mateus H. Gouveia, Marla Mendes, Gonçalo R. Abecasis, Isabela Alvim, Carlos E. Arboleda-Bustos, Gonzalo Arboleda, Humberto Arboleda, Mauricio L. Barreto, Lucas Barwick, Marcos A. Bezzera, John Blangero, Vanderci Borges, Omar Caceres, Jianwen Cai, Pedro Chana-Cuevas, Zhanghua Chen, Brian Custer, Michael Dean, Carla Dinardo, Igor Domingos, Ravindranath Duggirala, Elena Dieguez, Willian Fernandez, Henrique B. Ferraz, Frank D. Gilliland, Heinner Guio, Bernardo Horta, Joanne E. Curran, Jill M. Johnsen, Robert C. Kaplan, Shannon Kelly, Eimear E. Kenny, Barbara A. Konkle, Charles Kooperberg, Andres Lescano, M. Fernanda Lima-Costa, Ruth J. F. Loos, Ani Manichaikul, Deborah A. Meyers, Michel S. Naslavsky, Deborah A. Nickerson, Kari E. North, Carlos Padilla, Michael Preuss, Victor Raggio, Alexander P. Reiner, Stephen S. Rich, Carlos R. Rieder, Michiel Rienstra, Jerome I. Rotter, Tatjana Rundek, Ralph L. Sacco, Cesar Sanchez, Vijay G. Sankaran, Bruno Lopes Santos-Lobato, Artur Francisco Schumacher-Schuh, Marilia O. Scliar, Edwin K. Silverman, Tamar Sofer, Jessica Lasky-Su, Vitor Tumas, Scott T. Weiss, Latin American Research Consortium on the Genetics of Parkinson’s Disease (LARGE-PD), NINDS Stroke Genetics Network (SiGN) Consortium, TOPMed Population Genetics Working Group, Ignacio F. Mata, Ryan D. Hernandez, Eduardo Tarazona-Santos, Timothy D. O’Connor. Genetics of Latin American Diversity (GLAD) Project: insights into population genetics and association studies in recently admixed groups in the Americas. bioRxiv 2023.01.07.522490; doi: 10.1101/2023.01.07.522490

22. Harris DN, Song W, Shetty AC, Levano KS, Cáceres O, Padilla C, Borda V, Tarazona D, Trujillo O, Sanchez C, Kessler MD, Galarza M, Capristano S, Montejo H, Flores-Villanueva PO, Tarazona-Santos E, O’Connor TD, Guio H. Evolutionary genomic dynamics of Peruvians before, during, and after the Inca Empire. Proc Natl Acad Sci U S A. 2018 Jul 10;115(28):E6526–E6535. doi: 10.1073/pnas.1720798115. Epub 2018 Jun 26. PMID: 29946025; PMCID: PMC6048481

23. Jaramillo-Valverde L, Levano KS, Tarazona DD, Capristano S, Sanchez C, Poterico JA, Tarazona-Santos E, Guio H. Pharmacogenetic variability of tuberculosis biomarkers in native and mestizo Peruvian populations. Pharmacol Res Perspect. 2024 Jun;12(3):e1179. doi: 10.1002/prp2.1179. PMID: 38666760; PMCID: PMC11047445.

24. Levano KS, Jaramillo-Valverde L, Tarazona DD, Sanchez C, Capristano S, Vásquez-Loarte T, Solari L, Mendoza-Ticona A, Soto A, Rojas C, Zegarra-Chapoñan R, Guio H. Allelic and genotypic frequencies of NAT2, CYP2E1, and AADAC genes in a cohort of Peruvian tuberculosis patients. Mol Genet Genomic Med. 2021 Oct;9(10):e1764. doi: 10.1002/mgg3.1764. Epub 2021 Sep 12. PMID: 34510815; PMCID: PMC8580101.

25. Jaramillo-Valverde L, Levano KS, Tarazona DD, Capristano S, Zegarra-Chapoñan R, Sanchez C, Yufra-Picardo VM, Tarazona-Santos E, Ugarte-Gil C, Guio H. NAT2 and CYP2E1 polymorphisms and antituberculosis drug-induced hepatotoxicity in Peruvian patients. Mol Genet Genomic Med. 2022 Aug;10(8):e1987. doi: 10.1002/mgg3.1987. Epub 2022 Jun 24. PMID: 35751408; PMCID: PMC9356556.

26. Jaramillo-Valverde L, Levano KS, Tarazona DD, Vasquez-Dominguez A, Toledo-Nauto A, Capristano S, Sanchez C, Tarazona-Santos E, Ugarte-Gil C, Guio H. GSTT1/GSTM1 Genotype and Anti-Tuberculosis Drug-Induced Hepatotoxicity in Peruvian Patients. Int J Mol Sci. 2022 Sep 20;23(19):11028. doi: 10.3390/ijms231911028. PMID: 36232322; PMCID: PMC9569635.

27. Moreira RG, Saraiva-Duarte JM, Pereira AC, Sosa-Macias M, Galaviz-Hernandez C, Santolalla ML, Magalhães WCS, Zolini C, Leal TP, Balázs Z, Llerena A, Gilman RH, Mill JG, Borda V, Guio H, O’Connor TD, Tarazona-Santos E, Rodrigues-Soares F. Population genetics of PDE4B (phosphodiesterase-4B) in neglected Native Americans: Implications for cancer pharmacogenetics. Clin Transl Sci. 2022 Jun;15(6):1400–1405. doi: 10.1111/cts.13266. Epub 2022 Mar 28. PMID: 35266293; PMCID: PMC9199872.

28. Ganachari M, Guio H, Zhao N, Flores-Villanueva PO. Host gene-encoded severe lung TB: from genes to the potential pathways. Genes Immun. 2012 Dec;13(8):605–20. doi: 10.1038/gene.2012.39. Epub 2012 Sep 20. PMID: 22992722; PMCID: PMC3518758.

29. Vásquez-Loarte, T., Trubnykova, M. & Guio, H. Genetic association meta-analysis: a new classification to assess ethnicity using the association of MCP-1 – 2518 polymorphism and tuberculosis susceptibility as a model. BMC Genet 16, 128 (2015). 10.1186/s12863-015-0280-2

30. Yareta J, Galarza M, Capristano S, Pellón O, Ballon J, Guio H. Differential expression of circulating micro-RNAS in patients with active and latent tuberculosis. Rev Peru Med Exp Salud Publica. 2020;37(1): 51–6. Doi: 10.17843/rpmesp.2020.371.4468.

31. Guio H, Aliaga-Tobar V, Galarza M, Pellon-Cardenas O, Capristano S, Gomez HL, Olivera M, Sanchez C, Maracaja-Coutinho V. Comparative Profiling of Circulating Exosomal Small RNAs Derived From Peruvian Patients With Tuberculosis and Pulmonary Adenocarcinoma. Front Cell Infect Microbiol. 2022 Jun 30;12:909837. doi: 10.3389/fcimb.2022.909837. PMID: 35846752; PMCID: PMC9280157.

32. Jaramillo-Valverde L, Vasquez-Dominguez A, Levano KS, Novoa-Bellota P, Machacuay-Romero M, Garcia-de-la-Guarda R, Castrejon-Cabanillas R, Palma-Lozano D, Ashok KS, Davison S, Flores-Villanueva P, Cano RJ, Gomez A, Shady-Solis R, Guio H. Analysis of microbiome diversity in coprolites from Caral, Peru. Bioinformation. 2022 Dec 31;18(12):1159–1165. doi: 10.6026/973206300181159. PMID: 37701514; PMCID: PMC10492910.

33. Jaramillo-Valverde L, Vásquez-Domínguez A, Levano KS, Castrejon-Cabanillas R, Novoa-Bellota P, Machacuay-Romero M, Garcia-de-la-Guarda R, Cano RJ, Shady-Solis R, Guio H. A mobile lab for ancient DNA extraction in Peru. Bioinformation. 2022 Dec 31;18(12):1114–1118. doi: 10.6026/973206300181114. PMID: 37701515; PMCID: PMC10492912

34. Borda V, Alvim I, Mendes M, Silva-Carvalho C, Soares-Souza GB, Leal TP, Furlan V, Scliar MO, Zamudio R, Zolini C, Araújo GS, Luizon MR, Padilla C, Cáceres O, Levano K, Sánchez C, Trujillo O, Flores-Villanueva PO, Dean M, Fuselli S, Machado M, Romero PE, Tassi F, Yeager M, O’Connor TD, Gilman RH, Tarazona-Santos E, Guio H. The genetic structure and adaptation of Andean highlanders and Amazonians are influenced by the interplay between geography and culture. Proc Natl Acad Sci U S A. 2020 Dec 22;117(51):32557–32565. doi: 10.1073/pnas.2013773117. Epub 2020 Dec 4. PMID: 33277433; PMCID: PMC7768732.

35. Luis Jaramillo-Valverde, Gilderlanio Santana de Araújo, Julio A. Poterico, Catalina Martinez-Jaramillo, Vicky Roa-Linares, Sandra Alvites-Arrieta, Nelis Pablo-Ramirez, Milward Ubillus, Diana Palma-Lozano, Carolina Silva-Carvalho, Luca Vasconcelos-da-Gama, Lucas F Costa, Eduardo Tarazona-Santos, Soumya Raychaudhuri, Heinner Guio. The effect of altitude on the expression of immune-related genes in Peruvian rural indigenous. bioRxiv preprint doi: 10.1101/2024.03.06.583674

36. Poterico, J.A.; Jaramillo-Valverde, L.; Pablo-Ramirez, N.; Roa-Linares, V.C.; Martinez-Jaramillo, C.; Alvites-Arrieta, S.; Ubillus, M.; Palma-Lozano, D.; Castrejon-Cabanillas, R.; Davison, S.;, et al. Uncovering the Resistome of a Peruvian City through a Metagenomic Analysis of Sewage Samples. Environments 2023, 10, 191. 10.3390/environments10110191

37. Accinelli RA, Leon-Abarca JA. At High Altitude COVID-19 Is Less Frequent: The Experience of Peru. Arch Bronconeumol (Engl Ed). 2020 Nov;56(11):760–761. English, Spanish. doi: 10.1016/j.arbres.2020.06.015. Epub 2020 Jul 16. PMID: 32782091; PMCID: PMC7365056.

38. Alexander DH, Novembre J, Lange K. Fast model-based estimation of ancestry in unrelated individuals. Genome Res. 2009;19(9):1655–1664. doi:10.1101/gr.094052.109

39. Deurenberg P, Deurenberg-Yap M, Guricci S. Asians are different from Caucasians and from each other in their body mass index/body fat per cent relationship. Obes Rev. 2002 Aug;3(3):141–6. doi: 10.1046/j.1467-789x.2002.00065.x. PMID: 12164465.

40. Guimarães Alves AC, Sukow NM, Adelman Cipolla G, Mendes M, Leal TP, Petzl-Erler ML, Lehtonen Rodrigues Souza R, Rainha de Souza I, Sanchez C, Santolalla M, Loesch D, Dean M, Machado M, Moon JY, Kaplan R, North KE, Weiss S, Barreto ML, Lima-Costa MF, Guio H, Cáceres O, Padilla C, Tarazona-Santos E, Mata IF, Dieguez E, Raggio V, Lescano A, Tumas V, Borges V, Ferraz HB, Rieder CR, Schumacher-Schuh A, Santos-Lobato BL, Chana-Cuevas P, Fernandez W, Arboleda G, Arboleda H, Arboleda-Bustos CE, O’Connor TD, Beltrame MH, Borda V. Tracing the Distribution of European Lactase Persistence Genotypes Along the Americas. Front Genet. 2021 Sep 22;12:671079. doi: 10.3389/fgene.2021.671079. PMID: 34630506; PMCID: PMC8493957.

41. Corpas M, Siddiqui MK, Soremekun O, Mathur R, Gill D, Fatumo S. Addressing Ancestry and Sex Bias in Pharmacogenomics. Annu Rev Pharmacol Toxicol. 2024 Jan 23;64:53–64. doi: 10.1146/annurev-pharmtox-030823-111731. Epub 2023 Jul 14. PMID: 37450899.

42. 1000 Genomes Project Consortium; Auton A, Brooks LD, Durbin RM, Garrison EP, Kang HM, Korbel JO, Marchini JL, McCarthy S, McVean GA, Abecasis GR. A global reference for human genetic variation. Nature. 2015 Oct 1;526(7571):68–74. doi: 10.1038/nature15393. PMID: 26432245; PMCID: PMC4750478.

43. Yang HC, Chen CW, Lin YT, Chu SK. Genetic ancestry plays a central role in population pharmacogenomics. Commun Biol. 2021 Feb 5;4(1):171. doi: 10.1038/s42003-021-01681-6.

44. Nakatsuka N, Lazaridis I, Barbieri C, Skoglund P, Rohland N, Mallick S, Posth C, Harkins-Kinkaid K, Ferry M, Harney É, Michel M, Stewardson K, Novak-Forst J, Capriles JM, Durruty MA, Álvarez KA, Beresford-Jones D, Burger R, Cadwallader L, Fujita R, Isla J, Lau G, Aguirre CL, LeBlanc S, Maldonado SC, Meddens F, Messineo PG, Culleton BJ, Harper TK, Quilter J, Politis G, Rademaker K, Reindel M, Rivera M, Salazar L, Sandoval JR, Santoro CM, Scheifler N, Standen V, Barreto MI, Espinoza IF, Tomasto-Cagigao E, Valverde G, Kennett DJ, Cooper A, Krause J, Haak W, Llamas B, Reich D, Fehren-Schmitz L. A Paleogenomic Reconstruction of the Deep Population History of the Andes. Cell. 2020 May 28;181(5):1131–1145.e21. doi: 10.1016/j.cell.2020.04.015. Epub 2020 May 7. PMID: 32386546; PMCID: PMC7304944.

45. Arango-Isaza E, Capodiferro MR, Aninao MJ, Babiker H, Aeschbacher S, Achilli A, Posth C, Campbell R, Martínez FI, Heggarty P, Sadowsky S, Shimizu KK, Barbieri C. The genetic history of the Southern Andes from present-day Mapuche ancestry. Curr Biol. 2023 Jul 10;33(13):2602–2615.e5. doi: 10.1016/j.cub.2023.05.013. Epub 2023 Jun 5. PMID: 37279753.

46. Yang HC, Chen CW, Lin YT, Chu SK. Genetic ancestry plays a central role in population pharmacogenomics. Commun Biol. 2021 Feb 5;4(1):171. doi: 10.1038/s42003-021-01681-6.

47. Sharma V, Varshney R, Sethy NK. Human adaptation to high altitude: a review of convergence between genomic and proteomic signatures. Hum Genomics. 2022 Jul 15;16(1):21. doi: 10.1186/s40246-022-00395-y. PMID: 35841113; PMCID: PMC9287971.

48. Wang YF, Zhang Y, Lin Z, Zhang H, Wang TY, Cao Y, Morris DL, Sheng Y, Yin X, Zhong SL, Gu X, Lei Y, He J, Wu Q, Shen JJ, Yang J, Lam TH, Lin JH, Mai ZM, Guo M, Tang Y, Chen Y, Song Q, Ban B, Mok CC, Cui Y, Lu L, Shen N, Sham PC, Lau CS, Smith DK, Vyse TJ, Zhang X, Lau YL, Yang W. Identification of 38 novel loci for systemic lupus erythematosus and genetic heterogeneity between ancestral groups. Nat Commun. 2021 Feb 3;12(1):772. doi: 10.1038/s41467-021-21049-y. PMID: 33536424; PMCID: PMC7858632.

